# The pandemic, COVID-19 disease and perinatal health

**DOI:** 10.1101/2024.01.11.24301106

**Authors:** Shelley Jung, Emily F Liu, Dana E Goin, Kara E Rudolph, Mahasin S Mujahid, William H Dow, Jennifer Ahern

## Abstract

Adverse effects of COVID-19 on perinatal health have been documented, however there is a lack of research that separates individual disease from other changing risks during the pandemic period. We linked California statewide birth and hospital discharge data for 2019-2020, and compared health indicators among 3 groups of pregnancies: [a] 2020 delivery with COVID-19, [b] 2020 delivery with no documented COVID-19, and [c] 2019 pre-pandemic delivery. We aimed to quantify the links between COVID-19 and perinatal health, separating individual COVID-19 disease (a vs b) from the pandemic period (b vs c). We examined the following health indicators: preterm birth, hypertensive disorders of pregnancy, gestational diabetes mellitus and severe maternal morbidity. We applied model based standardization to estimate “average effect of treatment on the treated” risk differences (RD), and adjusted for individual and community-level confounders. Among pregnancies in 2020, those with COVID-19 disease had higher burdens of preterm birth (RD[95% confidence interval (CI)]=2.8%[2.1,3.5]), hypertension (RD[95% CI]=3.3%[2.4,4.1]), and severe maternal morbidity (RD[95% CI]=2.3%[1.9,2.7]) compared with pregnancies without COVID-19 (a vs b) adjusted for confounders. Pregnancies in 2020 without COVID-19 had a lower burden of preterm birth (RD[95% CI]=-0.4%[-0.6,-0.3]), particularly spontaneous preterm, and a higher burden of hypertension (RD[95% CI]=1.0%[0.9,1.2]) and diabetes RD[95%CI]=0.9%[0.8,1.1] compared with pregnancies in 2019 (b vs c) adjusted for confounders. Protective associations of the pandemic period for spontaneous preterm birth may be explained by socioenvironmental and behavioral modifications, while increased maternal conditions may be due to stress and other behavioral changes. To our knowledge, our study is the first to distinguish between individual COVID-19 disease and the pandemic period in connection with perinatal outcomes.

## INTRODUCTION

SARS-CoV-2 infection has physiologic effects on pregnancy, including through immune and inflammatory mechanisms that impact placental function, influencing perinatal health (1). To date, connections of COVID-19 with adverse perinatal outcomes have been documented in large population studies and meta-analyses (2). Among these studies, few have considered links of the pandemic period with perinatal health, and there is a particular lack of research that separates individual COVID-19 disease from other changing risks during the pandemic period. We aim to quantify the population-level associations of COVID-19 with birth parent and infant health, distinguishing the pandemic period from individual COVID-19 disease, to improve understanding of how the pandemic has impacted perinatal health.

## METHODS

We examined statewide California data, individually linking all birth and hospital discharge records for 2019-2020, subset to April to December in each year for seasonal comparability. We compared health indicators for mothers/birth parents and infants among 3 groups of pregnancies: [a] 2020 delivery with COVID-19 disease, [b] 2020 delivery with no documented COVID-19 disease, and [c] a 2019 pre-pandemic delivery. We examined the following health indicators: preterm birth, hypertensive disorders of pregnancy (HDP), gestational diabetes mellitus (GDM) and severe maternal morbidity (SMM). We separated COVID-19 disease (comparing [a] to [b]) from the broader societal changes during the pandemic period (comparing [b] to [c]). We applied model based standardization (g-computation)(3) to estimate a risk difference (RD) known as the ‘average effect of treatment on the treated’ (ATT), adjusted for available individual (insurance, parity, education, age, race and ethnicity) and community-level confounders (racial/ethnic composition, poverty, median household income, unemployment, vehicle ownership, vacancy rates, and marital status). We constructed 95% confidence intervals from 500 bootstrapped replications.

## RESULTS

The distribution of births and covariates by exposure group is in TABLE 1. The adjusted associations of COVID-19 disease at delivery are shown in TABLE 2a. There were higher burdens of preterm birth (risk difference (RD), [95% confidence interval (CI)]=2.8%[2.1, 3.5]), HDP (RD[95% CI]=3.3%[2.4, 4.1]), and SMM (RD[95% CI]=2.3%[1.9, 2.7]) among those with COVID-19 compared to 2020 pregnancies without COVID-19. The adjusted associations of the pandemic period are shown in TABLE 2b. There was a lower burden of preterm birth (RD[95% CI]=-0.4%[-0.6, -0.3]), particularly spontaneous preterm, and a higher burden of HDP (RD[95% CI]=1.1%[0.9, 1.2]) and GDM RD[95%CI]=0.9%[0.8, 1.1] among 2020 deliveries without COVID-19, compared with deliveries in 2019. Overall, COVID-19 disease was associated with adverse health for mothers/birth parents and infants. The pandemic period had smaller but meaningful protective associations with preterm birth and harmful associations with maternal conditions.

**TABLE 1.**
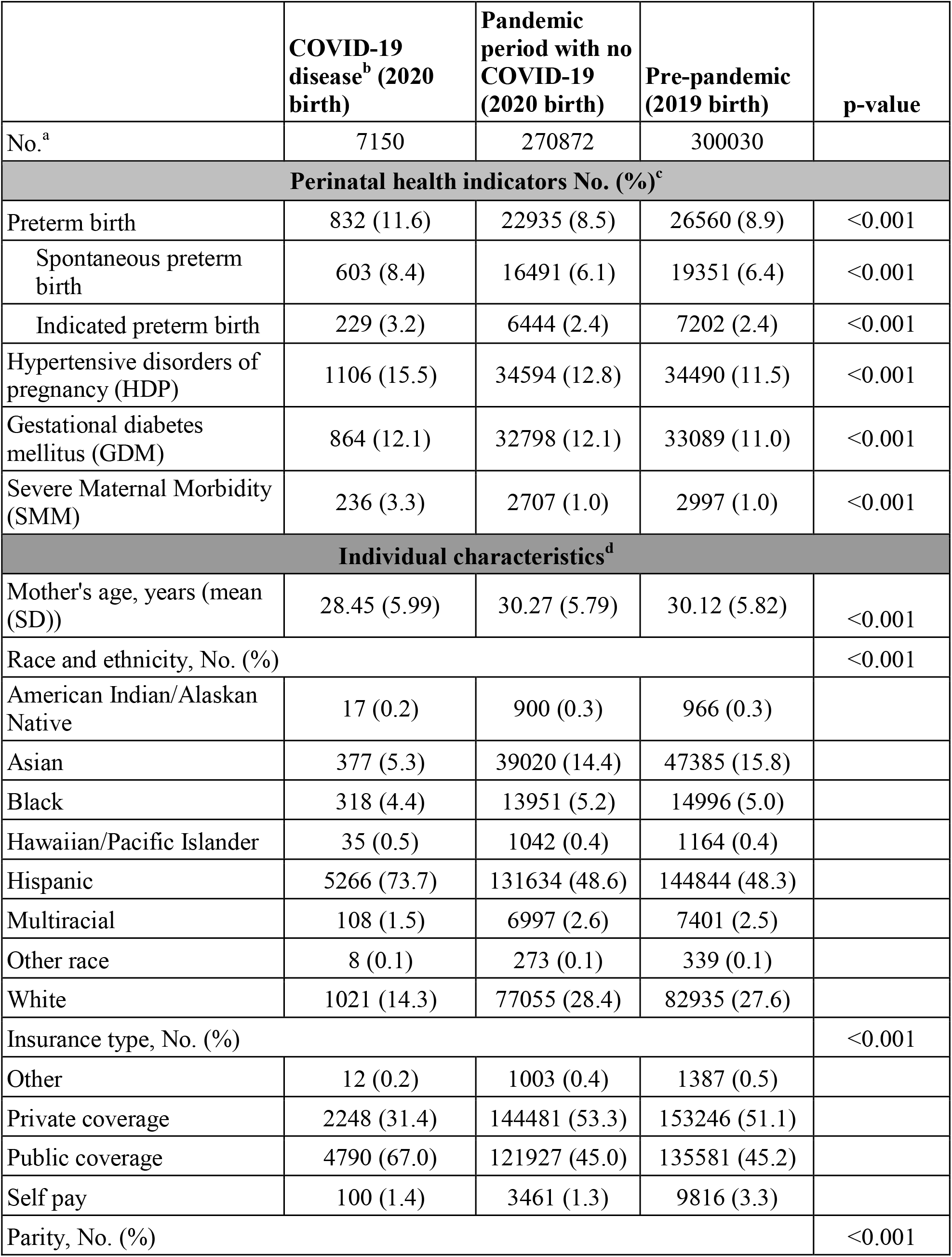
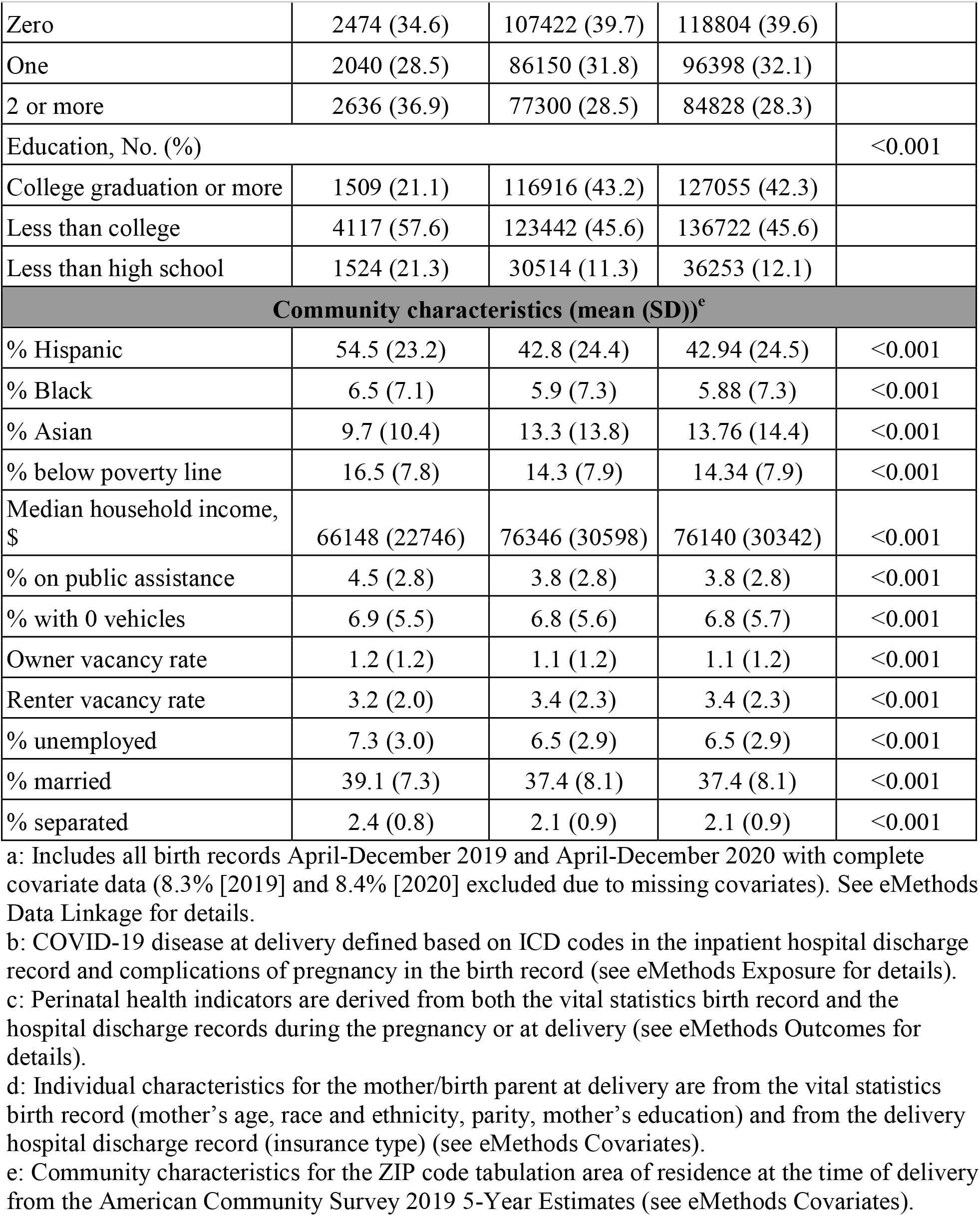
Characteristics of deliveries by COVID-19 disease and year.

|  | **COVID-19**<br>**disease<sup>b</sup> (2020**<br>**birth)** | **Pandemic**<br>**period with no**<br>**COVID-19**<br>**(2020 birth)** | **Pre-pandemic**<br>**(2019 birth)** | **p-value** |
| --- | --- | --- | --- | --- |
| No.<sup>a</sup> | 7150 | 270872 | 300030 |  |
| **Perinatal health indicators No. (%)<sup>c</sup>** |  |  |  |  |
| Preterm birth | 832 (11.6) | 22935 (8.5) | 26560 (8.9) | <0.001 |
| Spontaneous preterm birth | 603 (8.4) | 16491 (6.1) | 19351 (6.4) | <0.001 |
| Indicated preterm birth | 229 (3.2) | 6444 (2.4) | 7202 (2.4) | <0.001 |
| Hypertensive disorders of pregnancy (HDP) | 1106 (15.5) | 34594 (12.8) | 34490 (11.5) | <0.001 |
| Gestational diabetes mellitus (GDM) | 864 (12.1) | 32798 (12.1) | 33089 (11.0) | <0.001 |
| Severe Maternal Morbidity (SMM) | 236 (3.3) | 2707 (1.0) | 2997 (1.0) | <0.001 |
| **Individual characteristics<sup>d</sup>** |  |  |  |  |
| Mother's age, years (mean (SD)) | 28.45 (5.99) | 30.27 (5.79) | 30.12 (5.82) | <0.001 |
| Race and ethnicity, No. (%) |  |  |  | <0.001 |
| American Indian/Alaskan Native | 17 (0.2) | 900 (0.3) | 966 (0.3) |  |
| Asian | 377 (5.3) | 39020 (14.4) | 47385 (15.8) |  |
| Black | 318 (4.4) | 13951 (5.2) | 14996 (5.0) |  |
| Hawaiian/Pacific Islander | 35 (0.5) | 1042 (0.4) | 1164 (0.4) |  |
| Hispanic | 5266 (73.7) | 131634 (48.6) | 144844 (48.3) |  |
| Multiracial | 108 (1.5) | 6997 (2.6) | 7401 (2.5) |  |
| Other race | 8 (0.1) | 273 (0.1) | 339 (0.1) |  |
| White | 1021 (14.3) | 77055 (28.4) | 82935 (27.6) |  |
| Insurance type, No. (%) |  |  |  | <0.001 |
| Other | 12 (0.2) | 1003 (0.4) | 1387 (0.5) |  |
| Private coverage | 2248 (31.4) | 144481 (53.3) | 153246 (51.1) |  |
| Public coverage | 4790 (67.0) | 121927 (45.0) | 135581 (45.2) |  |
| Self pay | 100 (1.4) | 3461 (1.3) | 9816 (3.3) |  |
| Parity, No. (%) |  |  |  | <0.001 |
| Zero | 2474 (34.6) | 107422 (39.7) | 118804 (39.6) |  |
| One | 2040 (28.5) | 86150 (31.8) | 96398 (32.1) |  |
| 2 or more | 2636 (36.9) | 77300 (28.5) | 84828 (28.3) |  |
| Education, No. (%) |  |  |  | <0.001 |
| College graduation or more | 1509 (21.1) | 116916 (43.2) | 127055 (42.3) |  |
| Less than college | 4117 (57.6) | 123442 (45.6) | 136722 (45.6) |  |
| Less than high school | 1524 (21.3) | 30514 (11.3) | 36253 (12.1) |  |
| **Community characteristics (mean (SD))<sup>e</sup>** |  |  |  |  |
| % Hispanic | 54.5 (23.2) | 42.8 (24.4) | 42.94 (24.5) | <0.001 |
| % Black | 6.5 (7.1) | 5.9 (7.3) | 5.88 (7.3) | <0.001 |
| % Asian | 9.7 (10.4) | 13.3 (13.8) | 13.76 (14.4) | <0.001 |
| % below poverty line | 16.5 (7.8) | 14.3 (7.9) | 14.34 (7.9) | <0.001 |
| Median household income, \$ | 66148 (22746) | 76346 (30598) | 76140 (30342) | <0.001 |
| % on public assistance | 4.5 (2.8) | 3.8 (2.8) | 3.8 (2.8) | <0.001 |
| % with 0 vehicles | 6.9 (5.5) | 6.8 (5.6) | 6.8 (5.7) | <0.001 |
| Owner vacancy rate | 1.2 (1.2) | 1.1 (1.2) | 1.1 (1.2) | <0.001 |
| Renter vacancy rate | 3.2 (2.0) | 3.4 (2.3) | 3.4 (2.3) | <0.001 |
| % unemployed | 7.3 (3.0) | 6.5 (2.9) | 6.5 (2.9) | <0.001 |
| % married | 39.1 (7.3) | 37.4 (8.1) | 37.4 (8.1) | <0.001 |
| % separated | 2.4 (0.8) | 2.1 (0.9) | 2.1 (0.9) | <0.001 |

a: Includes all birth records April-December 2019 and April-December 2020 with complete covariate data (8.3% [2019] and 8.4% [2020] excluded due to missing covariates). See eMethods Data Linkage for details.
b: COVID-19 disease at delivery defined based on ICD codes in the inpatient hospital discharge record and complications of pregnancy in the birth record (see eMethods Exposure for details).
c: Perinatal health indicators are derived from both the vital statistics birth record and the hospital discharge records during the pregnancy or at delivery (see eMethods Outcomes for details).
d: Individual characteristics for the mother/birth parent at delivery are from the vital statistics birth record (mother's age, race and ethnicity, parity, mother's education) and from the delivery hospital discharge record (insurance type) (see eMethods Covariates).
e: Community characteristics for the ZIP code tabulation area of residence at the time of delivery from the American Community Survey 2019 5-Year Estimates (see eMethods Covariates).

**TABLE 2.** Adjusted marginal risk differences for (a) COVID-19 disease, and (b) the pandemic period.

| **(a) COVID-19 at delivery (2020, No.=7150) vs pandemic period with no COVID-19 at delivery (2020, No.=270,872)<sup>a</sup>** |  |  |  |  |
| --- | --- | --- | --- | --- |
| **Outcome** | **R1<sup>b</sup>** | **R0<sup>c</sup>** | **ATT (RD)<sup>d</sup>** | **95% CI** |
| Preterm birth | 0.116 | 0.089 | 0.028 | (0.021, 0.035) |
| Spontaneous preterm birth | 0.084 | 0.064 | 0.020 | (0.014, 0.027) |
| Indicated preterm birth | 0.032 | 0.025 | 0.007 | (0.004, 0.011) |
| HDP | 0.155 | 0.122 | 0.033 | (0.024, 0.041) |
| GDM | 0.121 | 0.114 | 0.007 | (0.000, 0.015) |
| SMM | 0.033 | 0.010 | 0.023 | (0.019, 0.027) |
| **(b) Pandemic period with no COVID-19 (2020, No.=270,872) vs pre-pandemic delivery (2019, No.=300,030)<sup>a</sup>** |  |  |  |  |
| Preterm birth | 0.085 | 0.089 | -0.004 | (-0.006, -0.003) |
| Spontaneous preterm birth | 0.061 | 0.065 | -0.004 | (-0.005, -0.003) |
| Indicated preterm birth | 0.024 | 0.024 | 0.000 | (-0.001, 0.000) |
| HDP | 0.128 | 0.117 | 0.011 | (0.009, 0.012) |
| GDM | 0.121 | 0.112 | 0.009 | (0.008, 0.011) |
| SMM | 0.010 | 0.010 | 0.000 | (-0.001, 0.000)) |

b: R1 is the estimated risk of the outcome if the exposed group (e.g., group with COVID-19 disease) experienced the exposure, adjusted for covariates.
c: R0 is the estimated risk of the outcome if the exposed group had not experienced the exposure, adjusted for covariates.
d: ATT (RD) is the risk difference R1-R0, that estimates the amount of additional risk experienced by the exposed group (e.g., group with COVID-19 disease, or pandemic period deliveries) that is due to the exposure.

## DISCUSSION

This research adds to the understanding of the overall impacts of COVID-19 on perinatal health in a large, diverse general population sample of the US by distinguishing the connections of COVID-19 disease from those of the pandemic period with perinatal health. Our findings suggest that COVID-19 disease was harmful for preterm birth, HDP, and SMM, consistent with other studies (2,4). Interestingly, we found the pandemic period decreased risk of preterm birth, but increased risk of HDP and GDM; this has been reported elsewhere, but not using designs that could disentangle infection from the pandemic period (5,6). The pandemic period protective association with preterm birth was largest for spontaneous preterm, suggesting that it may be explained by socioenvironmental and behavioral modifications, such as improved air quality, changes in commuting patterns, and reduced non-COVID infections. Increased risk of HDP and GDM may be due to stress, changes in eating patterns, and reduced or remote prenatal care due to the pandemic. To our knowledge, our study is the first to distinguish between individual COVID-19 disease and the pandemic period in connection with perinatal outcomes.

## Data Availability

Birth and hospital discharge data are available upon request for research projects from the California Department of Public Health, Center for Health Statistics and Informatics, and from the California Department of Health Care Access and Information.

