## Supplementary Methods for "The pandemic, COVID-19 disease and perinatal health"

### eMethods

1. Data Linkage
2. Exposure
3. Outcomes
4. Covariates
5. Analysis

**1. Data Linkage**

To create the linked data set, we worked with the 2019 Birth Cohort File and 2020 Comprehensive Birth File (released earlier than the Birth Cohort File) from the California Department of Public Health, Center for Health Statistics and Informatics, and the 2019 and 2020 inpatient hospital discharge data (Patient Discharge Data PDD) from the California Department of Health Care Access and Information. The primary approach was to link births with maternal hospital records on the hospital identification code and the mother's date of birth. If a match was not identified, mother's admit date, baby's date of birth and mother's ZIP code of residence were used to match. If a match was still not identified, mother's date of birth and mother's ZIP code of residence were used to match. Using the primary linkage approach, 96.4% and 96.2% of hospital births were matched in 2019 and 2020, respectively. Overall (after the primary and secondary approaches), 97.3% and 97.4% of hospital births were matched in 2019 and 2020, respectively.

**2. Exposure**

Deliveries were considered to have COVID-19 disease if the inpatient hospital discharge record included the ICD-10 code U071 for COVID-19 (introduced in April 2020), or if the birth record included the "Complications and procedures of pregnancy and concurrent illness" codes 91 and 92 for presumed and confirmed COVID-19, respectively (introduced in June 2020). Deliveries were considered exposed to the pandemic period if they took place April-December 2020, and did not have a record of COVID-19. Deliveries April-December 2019 were unexposed to SARS-CoV-2 infection and the pandemic period. Recent research on California deliveries during the pandemic ascertained that about 85% of births were in facilities with confirmed universal testing, and that associations between COVID-19 and outcomes were very similar when restricting to the universal testing subset of deliveries(1).

**3. Outcomes**

Perinatal health outcomes were classified based on combinations of the birth record codes and hospitalization ICD10 codes from pregnancy or delivery, as specified in eTable 1. Where ICD10 codes were used, the specific codes for each outcome are listed in eTable 2.

eTable 1. Definitions of perinatal health outcomes

| OUTCOME | DEFINITION |
|---|---|
| Preterm birth | Gestational weeks at birth less than 37 weeks (birth record) |
| Spontaneous (2) | Preterm premature rupture of membranes (PPROM) (ICD10 or birth record) |
| | Vaginal delivery without PPROM (ICD10 or birth record) |
| Indicated (2) | Preterm induction of labor (IOL) without PPROM (ICD10 or birth record) |
| | Cesarean delivery without IOL or PPROM (ICD10 or birth record) |
| Hypertensive disorder of pregnancy (HDP) (3) | Preeclampsia (ICD10 code or birth record) |
| | Eclampsia (ICD10 code or birth record) |
| | Hypertension during pregnancy (ICD10 or birth record) |
| | No record of prior hypertension (ICD10 or birth record) |
| Gestational diabetes mellitus (GDM) (4) | Gestational diabetes (ICD10 code or birth record) |
| | No record of prior diabetes (ICD10 code or birth record) |

| | |
|---|---|
| Severe maternal morbidity (SMM) (5) | SMM (ICD10 codes [without transfusion]) |

eTable 2. ICD10 codes from the hospitalization records used to identify elements of perinatal outcomes

| CONDITION | ICD10 CODE |
|---|---|
| Preterm premature rupture of membranes (2) | O42011, O42012, O42013, O42019, O42111, O42112, O42113, O42119, O42911, O42912, O42913, O42919, P011 |
| Preeclampsia (3) | O11, O14 |
| Eclampsia (3) | O15 |
| Hypertension during pregnancy (3) | O10, O11, O13, O14, O15, O16 |
| Pre-pregnancy hypertension (3) | I10, I11, I12, I13, I15 |
| Gestational diabetes (4) | O244, O249, O9981 |
| Diabetes (4) | E08, E09, E10, E11, E13, O240, O241, O243, O248 |
| SMM (diagnosis codes) (5) | I2101, I2102, I2109, I2111, I2119, I2121, I2129, I213, I214, I219, I21A1, I21A9, I220, I221, I222, I228, I229, I7100, I7101, I7102, I7103, I711, I712, I713, I714, I715, I716, I718, I719, I790, N170, N171, N172, N178, N179, O904, J80, J951, J952, J953, J95821, J95822, J9600, J9601, J9602, J9620, J9621, J9622, R092, O88111, O88112, O88113, O8812, O8813, I462, I468, I469, I4901, I4902, D65, D688, D689, O723, O1500, O1502, O1503, O151, O152, O159, O1422, O1423, I97120, I97121, I97130, I97131, I97710, I97711, I6000, I6001, I6002, I6010, I6011, I6012, I602, I6030, I6031, I6032, I604, I6050, I6051, I6052, I606, I607, I608, I609, I611, I612, I613, I614, I615, I616, I618, I619, I6200, I6201, I6202, I6203, I621, I629, I6300, I63011, I63012, I63013, I63019, I6302, I63031, I63032, I63033, I63039, I6309, I6310, I63111, I63112, I63113, I63119, I6312, I63131, I63132, I63133, I63139, I6319, I63211, I63212, I63213, I63219, I6322, I63231, I63232, I63233, I63239, I6329, I6330, I63311, I63312, I63313, I63319, I63321, I63322, I63323, I63329, I63331, I63332, I63333, I63339, I63341, I63342, I63343, I63349, I6339, I6340, I63411, I63412, I63413, I63419, I63421, I63422, I63423, I63429, I63431, I63432, I63433, I63439, I63441, I63442, I63443, I63449, I6349, I6350, I63511, I63512, I63513, I63519, I63521, I63522, I63523, I63529, I63531, I63532, I63533, I63539, I63541, I63542, I63543, I63549, I6359, I636, I6381, I6389, I639, I6501, I6502, I6503, I6509, I651, I6521, I6522, I6523, I6529, I658, I659, I6601, I6602, I6603, I6609, I6611, I6612, I6613, I6619, I6621, I6622, I6623, I6629, I663, I668, I669, I670, I671, I672, I673, I674, I675, I676, I677, I6781, I6782, I6783, I67841, I67848, I67850, I67858, I6789, I679, I680, I682, I688, O2251, O2252, O2253, I97810, I97811, I97820, I97821, O873, J810, I501, I5020, I5021, I5023, I5030, I5031, I5033, I5040, I5041, I5043, I509, O740, O741, O742, O743, O8901, O8909, O891, O892, O85, O8604, T80211A, T8140XA, T8141XA, T8142XA, T8143XA, T8144XA, T8149XA, T8144, T8144XD, T8144XS, R6520, A400, A401, A403, A408, A409, A4101, A4102, A411, A412, A413, A414, A4150, A4151, A4152, A4153, A4159, A4181, A4189, A419, A327, O751, R570, R571, R578, R579, R6521, T782XXA, T882XXA, T886XXA, T8110XA, T8111XA, T8119XA, D5700, D5701, D5702, D57211, D57212, D57219, D57411, D57412, D57419, D57811, D57812, D57819, I2601, I2602, I2609, I2690, I2692, I2699, O88011, O88012, O88013, O88019, O8802, O8803, O88211, O88212, O88213, O88219, O8822, O8823, O88311, O88312, O88313, O88319, O8832, O8833, O8881, O8882, O8883 |
| SMM (procedure codes) (5) | 5A2204Z, 5A12012, 0UT90ZZ, 0UT94ZZ, 0UT97ZZ, 0UT98ZZ, 0UT9FZZ, 0B110Z4, 0B110F4, 0B113Z4, 0B113F4, 0B114Z4, 0B114F4, 5A1935Z, 5A1945Z, 5A1955Z |

**4. Covariates**

We adjusted for individual and community-level demographic and socioeconomic characteristics that may confound the relation between COVID-19 disease or the pandemic period and the health outcomes of mothers/birth parents and infants. Individual characteristics for the mother/birth parent at delivery are from the vital statistics birth record (mother's age, race and ethnicity, parity, mother's education) and from the delivery hospital discharge record (insurance type). Community characteristics for the ZIP code tabulation area of residence at the time of delivery are from the American Community Survey 2019 5-Year Estimates. For COVID-19 disease, it is well documented that particular racial and ethnic, socioeconomic and age groups were at higher risk of COVID-19 disease due to jobs that could not be done remotely, increased crowding, and numerous other vulnerabilities to infection (6). Furthermore, community characteristics that capture structural inequities (such as segregation), or manifestations of those inequities, have been documented to increase the risk of COVID-19 beyond individual risk factors (6). These individual and community characteristics also have well established connections with gestational health (7,8). For the pandemic period, the characteristics of deliveries are less likely to be confounded because most 2020 deliveries were conceived before the pandemic, but control for these individual and community characteristics is a conservative approach to account for any small differences between the years.

**5. Analysis**

We used model-based standardization, also known as g-computation, for the analysis (9). First, a regression model (logistic regression) was used to estimate the relation between the exposure (COVID-19 disease, or the pandemic period) and outcome (each of the perinatal health outcomes), adjusting for confounders. Next, that model was used to impute the outcome for each observation under different exposures. In this application, we imputed outcomes while setting exposure to 1 (exposure to COVID-19 disease, or exposure to the pandemic period), and then imputed outcomes while setting exposure to 0 (no COVID-19 disease, or no pandemic period exposure). The imputed values were averaged across the population of interest, in this case they were averaged across the exposed group for each analysis in order to estimate the 'average effect of treatment on the treated' (ATT). We take the difference between two averaged risks to estimate the ATT: R1 - the estimated risk of the outcome if the exposed group (e.g., COVID-19 disease) experienced the exposure, adjusted for covariates, and R0 - the estimated risk of the outcome if the exposed group had not experienced the exposure, adjusted for covariates. ATT (RD) is the risk difference R1-R0, which estimates the amount of additional risk experienced by the exposed group (group with COVID-19 disease, or pandemic period delivery group) that is due to the exposure. Finally, we used the percentile method to construct 95% confidence intervals from 500 bootstrapped replications.
